# Systematic review of the clinical effectiveness of Tixagevimab/Cilgavimab for prophylaxis of COVID-19 in immunocompromised patients

**DOI:** 10.1101/2022.11.07.22281786

**Authors:** Rhea Suribhatla, Thomas Starkey, Maria C Ionescu, Antonio Pagliuca, Alex Richter, Lennard YW Lee

**Author notes:** **Corresponding Author:** Lennard YW Lee, Department of Oncology, University of Oxford, Oxford OX3 7DQ, 01865 617 331, Institute of Cancer and Genomic Sciences, University of Birmingham, Edgbaston, Birmingham B15 2TT, UK., 0121 414 3511.

## Abstract

**Background and aims:** Immunocompromised patients have a reduced ability to generate antibodies after COVID-19 vaccination and are at higher risk of SARS-CoV-2 infection, complications and mortality. Tixagevimab/Cilgavimab (Evusheld) is a monoclonal antibody combination which bind to the SARS-CoV-2 spike protein, preventing the virus entering human cells. The phase III PROVENT trial reported that immunocompromised patients given Tixagevimab/Cilgavimab had a significantly reduced risk of COVID-19 infection. However, PROVENT was conducted before the SARS-CoV-2 Omicron became prevalent. This systematic review provides an updated summary of real-world clinical evidence of Tixagevimab/Cilgavimab effectiveness in immunocompromised patients.

**Methods:** Two independent reviewers conducted PubMed and medRxiv searches for the period of 01/01/2021 to 01/10/2022. Clinical studies which reported the primary outcome of breakthrough COVID-19 infections after Tixagevimab/Cilgavimab administration were included in the review. COVID-19-related hospitalisations, ITU admissions and mortality were assessed as secondary outcomes. Clinical effectiveness was determined using the case-control clinical effectiveness methodology. The GRADE tool was used to ascertain the level of certainty for the primary outcome in each study.

**Results:** 17 clinical studies were included, comprising 24,773 immunocompromised participants of whom 10,775 received Tixagevimab/Cilgavimab. Most studies reported clinical outcomes during the SARS-CoV-2 Omicron wave. Six studies compared a Tixagevimab/Cilgavimab intervention group to a control group. Overall, the clinical effectiveness of prophylactic Tixagevimab/Cilgavimab against COVID-19 breakthrough infection, hospitalisation and ITU admission were 40.47%, 69.23% and 87.89%, respectively. For prevention of all-cause and COVID-19-specifc mortality, overall clinical effectiveness was 81.29% and 86.36%, respectively.

**Conclusions:** There is a growing body of real-world evidence validating the original PROVENT phase III study regarding the clinical effectiveness of Tixagevimab/Cilgavimab as prophylaxis for immunocompromised patients, notably demonstrating effectiveness during the Omicron wave. This review demonstrates the clinical effectiveness of prophylactic Tixagevimab/Cilgavimab at reducing COVID-19 infection, hospitalisation, ITU admission and mortality for immunosuppressed individuals. It is important that ongoing larger-scale and better-controlled real world studies are initiated and evaluated to provide ongoing certainty of the clinical benefit of prophylactic antibody treatment for immunocompromised patients in the face of new variants.

## Introduction

Since the start of the COVID-19 pandemic in March 2020, understanding of SARS-CoV-2 infection susceptibility and treatment has increased. Vaccination has been proven to protect individuals from the more serious effects of COVID-19, however, patients with weakened immune systems are unlikely to gain the same level of protection. Findings from the NIHR OCTAVE trial have revealed that, after two COVID-19 vaccinations, approximately 11% of immunocompromised patients did not generate any anti-COVID-19 antibodies after 4 weeks^1^

Patients may be immunocompromised for a number of medical reasons. This may be as a result of morbidities, such as cancer, blood cancer, solid organ and bone marrow transplant, renal disease, autoimmune disease, primary immunodeficiencies, liver disease, HIV or immune mediated inflammatory disorders, or because of treatments they are receiving immunosuppressants such as steroids, calcineurin inhibitors, disease modifying agents or chemotherapy. In the UK, there are around 500,000 immunocompromised people^2^. These individuals have been shown to be at significantly increased risk of COVID-19-related complications compared to the population^3–5^.

Tixagevimab/Cilgavimab, also known as Evusheld or AZD7442, is a combination of the two human monoclonal antibodies tixagevimab and cilgavimab, given as separate intramuscular injections within the same session. The antibodies bind to independent segments of the SARS-CoV-2 spike protein, preventing the virus binding to the human ACE2 receptor and entering human cells^6^. The antibodies are modified to increase relative to other monoclonal antibodies^6^. A prophylactic dosage of 600mg-300/300mg of tixagevimab/ cilgavimab is recommended^7^.

The phase III PROVENT trial was the first clinical study for Tixagevimab/Cilgavimab. The trial included adults who were unlikely to develop an adequate vaccination response and/or were at increased risk of SARS-CoV-2 exposure. The 3,460 patients prophylactically given Tixagevimab/Cilgavimab had a 76.6% reduced risk of COVID-19 infection after 3 months, and an 82.8% risk reduction after 6 months, compared to 1,737 control patients^8^. These results suggest that Tixagevimab/Cilgavimab could provide valuable protection for those most vulnerable to COVID-19. COVID-19 prophylaxis could reduce shielding restrictions for immunocompromised groups, potentially improving mental health and empowering a return to everyday life.

In March 2022, the Europeans Medicine Agency (EMA)^9^ and the United Kingdom Medicines and Healthcare products Regulatory Agency (MHRA) approved the use of Tixagevimab/Cilgavimab for adults who are unlikely to gain protection against Covid-19 from vaccination, or for whom vaccination is not recommended^10^. As of August 2022, Tixagevimab/Cilgavimab is in use in over 32 different countries, including the United States, France and Canada, Malaysia and Japan^11^.

However, the PROVENT study assessed Tixagevimab/Cilgavimab prior to the SARS-CoV-2 omicron variants. There is also doubt of the degree of clinical effectiveness in a real-world setting, particularly in preventing hospitalisations and deaths. Furthermore, some studies have cast doubt on the validity of prophylactic measures for immunocompromised patients^12^. Finally, there is a viewpoint that pandemic measures can be limited, curtailed or reallocated as a result of perceived decreased severity of the current variants^13^.

This systematic review provides an up-to-date summary of the clinical evidence of the efficacy of Tixagevimab/Cilgavimab in immunocompromised patients and aims to address some of the ongoing questions regarding the use of prophylactic antibody therapies with Tixagevimab/Cilgavimab for the immunocompromised population.

## Methods

### Study design

This was a systematic review of clinical studies in peer-reviewed journal articles and pre-print articles (Prospero registration 348513).

### Search strategy

Two independent reviewers conducted an electronic search strategy of two online databases, PubMed and medRxiv, including studies published in the period of 1^st^ January 2021 to 1^st^ October 2022. The initial search was performed with search terms including “Evusheld”, “tixagevimab”, “cilgavimab”, “AZD7442”, “AZD8895” and “AZD1061”. The reviewers then assessed each paper generated from the search and excluded articles firstly based on title, then abstract, then following review of the full text. References of the filtered papers were searched for additional studies. Additional studies were supplemented on the 1^st^ of October following the same search strategy at the request senior policy makers. Any disagreements between the reviewers were resolved by consulting a separate adjudicator and a discussion between all three parties.

### Eligibility and exclusion criteria

Eligible studies had to meet the following criteria: (1) involved a clinical study-randomised controlled trials, observational studies or cohort studies, (2) prophylaxis treatment with Tixagevimab/Cilgavimab, (3) report of breakthrough SARS-CoV-2 infections. Other descriptions of clinical outcomes, including hospitalisation, ITU admission or deaths, and sera SARS-CoV-2 neutralising ability were reported as secondary outcomes.

### Data extraction

Once all papers from the search had been identified the two independent reviewers reviewed the full text of all identified papers. Descriptive data for each article were identified including; study design-date accessed, title, year of publication, author, author contact details, funding sources; methodology-study aim, design, setting, start and end date, duration of participation; population-population description, setting, total number randomised, baseline imbalances, withdrawals and exclusions, age, sex (percent female), race/ ethnicity, severity of illness, co-morbidities, sociodemographic data, reason for being high risk for COVID-19; intervention-number randomised to group, description, duration of treatment period, timing, delivery method, providers, co-interventions, economic information, resource requirements, integrity of delivery; comparator(s)-number randomised to group, description, duration of treatment period, timing, delivery method, providers, co-interventions, economic information, resource requirements, integrity of delivery; outcome measures-number of COVID-19 infections, infection complications, hospitalisations, deaths and the neutralising ability of sera against SARS-CoV-19. For each outcome, the time point and duration, analysis scale and unit, number of missing participants and why, statistical methods used, and appropriateness was noted and Information for risk of bias and GRADE assessments.

Once all papers from the search had been identified the two independent reviewers reviewed the full text of all identified papers. Descriptive data for each article were identified including study design, methodology, population, intervention, comparator(s) and outcome measures. For each outcome, the time point and duration, analysis scale and unit, number of missing participants and why, statistical methods used, and appropriateness was noted and Information for risk of bias and GRADE assessments.

### Systematic review analysis

The risk of bias was conducted for each randomised controlled trial by two independent reviewers. A study is deemed at overall high risk if it is rated at high risk for any domain, at low risk if it is deemed low risk for all domains, and all other studies deemed at unclear risk. The GRADE tool was used to assess the certainty of the evidence for the primary outcome, judged against the following domains: participants follow up, imprecision, inconsistency, indirectness and publication bias^14^. GRADE levels were assigned as follows: (-)(-)(-)(-) = Level 0, (+)(-)(-)(-) = Level 1, (+)(+)(-)(-) = Level 2, (+)(+)(+)(-) = Level 3 and (+)(+)(+)(+) = Level 4.

### Data analysis

The outcomes assessed in this study included: COVID-19 infection (confirmed through any approved method of detecting COVID-19 infection including polymerase chain reaction tests (PCR) and lateral flow tests (LFTs)), admissions to hospital, including ICU, due to COVID-19 infection, mortality due to COVID-19 infection and all-cause mortality, and participant serum SARS-CoV-2 neutralising ability.

The clinical effectiveness of Tixagevimab/Cilgavimab was determined using the case-control clinical effectiveness methodology, similar to test-negative case control (TNCC) as previously reported for determining clinical effectiveness of SARS-CoV-2 vaccines^15^. Odds Ratios and 95% confidence intervals (CI) between intervention and control patient groups were calculated using a two-sided Fisher’s Exact test, subtracted from 1 and multiplied by 100 to calculate each clinical effectiveness percentage. Aggregated clinical effectiveness percentages of multiple studies were generated where corresponding intervention and control data was available^8,16–20^. 95% confidence intervals for infection and case-outcome percentages were calculated using the Wilson score method without continuity correction^21^. Statistical analyses and data visualisation were performed within R version 4.1.2.

## Results

The search strategy initially generated a total of 108 papers, 40 on medRxiv and 68 on PubMed. These were narrowed down to 17 full text articles based on the study inclusion criteria. The overall search strategy for the systematic review is set out in the PRISMA diagram (Figure 1). In total these seventeen studies included 24,773 immunocompromised participants, of whom 10,775 received Tixagevimab/Cilgavimab as a prophylactic antibody therapy. It included studies from the United States, France, Israel, Belgium, Spain and the United Kingdom.

**Figure 1:**
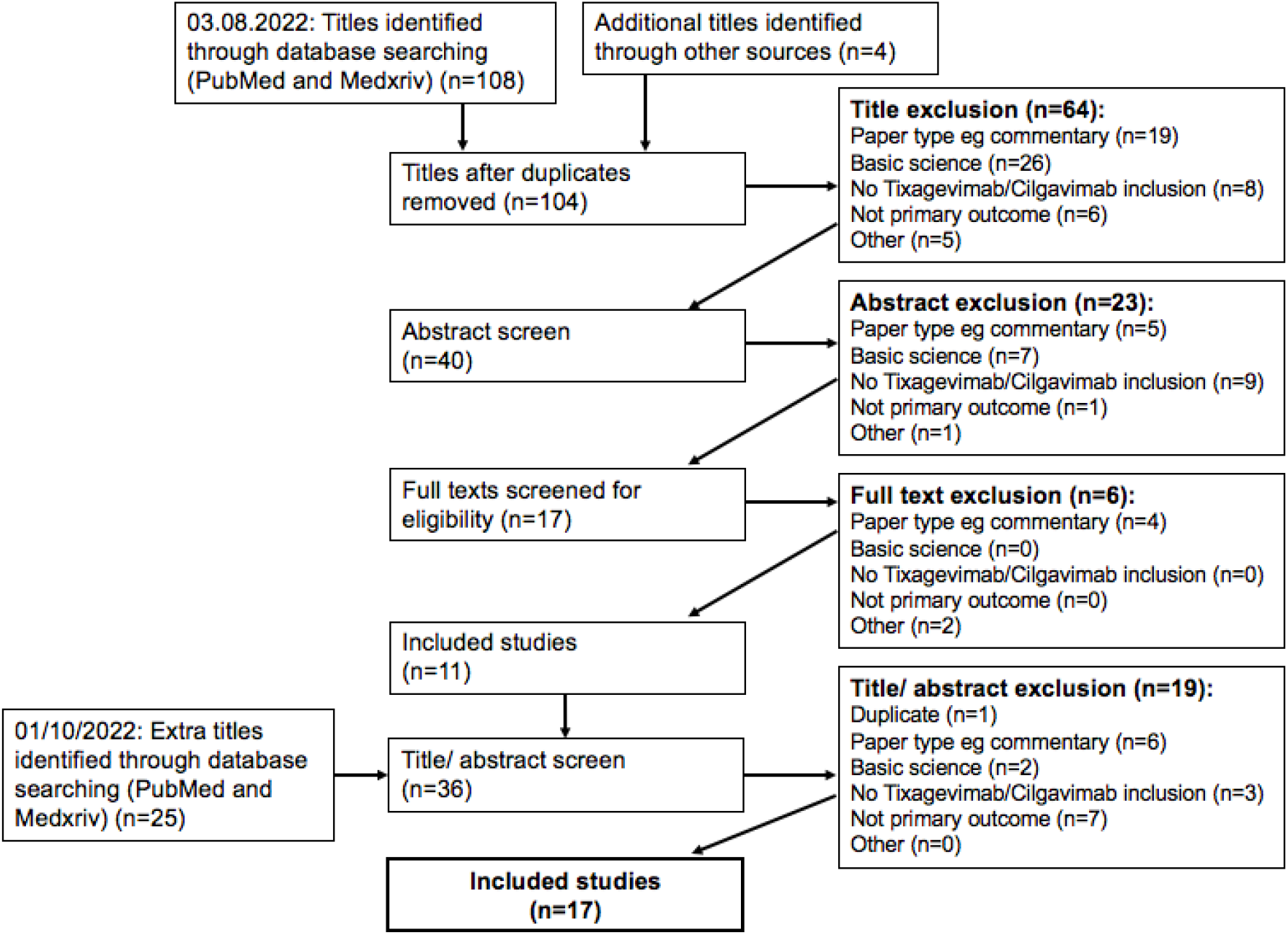
Prisma flowchart showing systematic processing of articles.

This review included ten retrospective cohort studies^12,16–20,22–25^ (including two preprints^16,20^), six prospective, observational cohort studies^26–31^ (also including one preprint^28^) and one randomised controlled trial^8^. The studies included adult patients who were immunocompromised or at high risk of COVID-19 infection. Patients were immunocompromised as a result of having a solid organ transplant, chemotherapy, receiving immunosuppressant drugs like steroids or anti-CD20s, or having a blood cancer. The majority of the studies reported on clinical outcomes during the Omicron wave. The findings from the papers are summarised in Figure 2 and Tables 1-3, with full details of study characteristics in the Appendix.

**Figure 2:**
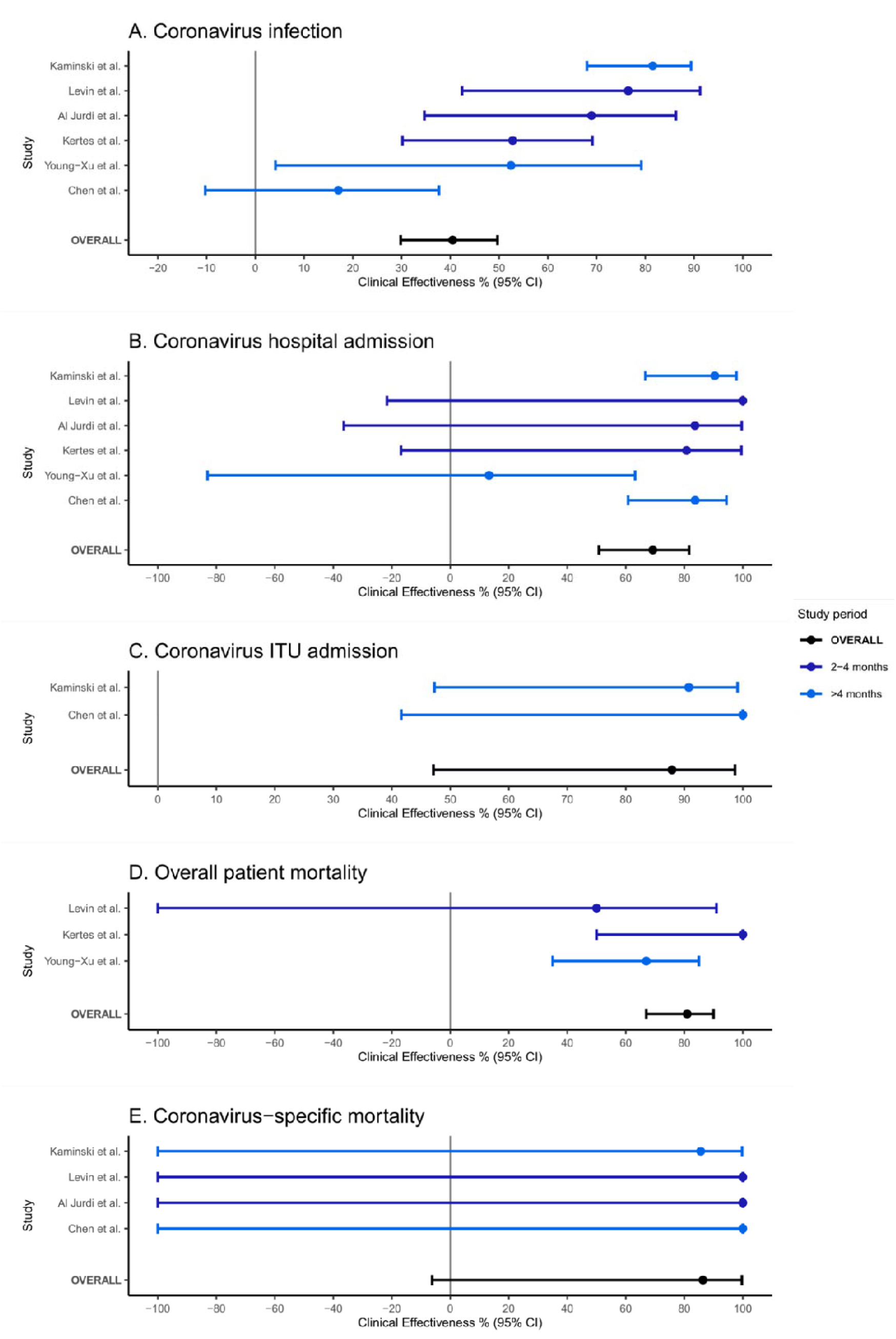
Box and whisker plot showing Clinical effectiveness of Tixagevimab/Cilgavimab against breakthrough coronavirus infection, hospitalisation, ITU admission, mortality and coronavirus-specific mortality ^8,12,16–20,22–31^

**Table 1:** Primary outcome summary ^8,12,16–20,22–31^. (RRR: relative risk ratio; HR: hazard ratio; IRR: incidence risk ratio; OR: odds ratio; IQR: interquartile range).

| Paper | Tixagevimab/Cilgavimab dosage | Infections<br>Infections/ no of participants<br>% (95% CI) |  | Relative effect<br>(95% CI) or<br>significance | Time |
| --- | --- | --- | --- | --- | --- |
|  |  | Intervention | Control |  |  |
| Levin et al | 150/150mg (100%) | 8/3441, 0.23% (0.12-0.46) | 17/1731, 0.98% (0.61-1.57) | RRR 76.7% (95% CI 46.0-90.0)<br>p<0.001 | Median of 83 days |
|  |  | 11/3441, 0.32% (0.18-0.57) | 31/1731, 1.79% (1.26-2.53) | RRR 82.8% (95% CI 65.8-91.4%) | Median of 196 days |
| Young-Xu et al. (preprint) | 150/150mg (17%),<br>300/300mg (83%) | <9/1733, 0.52% (0.27-0.98) | 69/6354, 1.09% (0.86-1.37) | HR 0.34 (0.13-0.87)<br>IRR 0.32 (0.24-0.44) | Over 4 months |
| Al Jurdi et al. | 150/150mg (40.5%),<br>300/300mg (59.0%),<br>450/450mg (0.5%) | 11/222, 4.95% (2.79-8.65) | 32/222, 14.41% (10.40-19.64) | Log-rank p<0.001 | Mean of 87 ± 30 days after Tixagevimab/Cilgavimab; mean of 82 ± 28 after control |
| Kertes et al. | 150/150mg (100%) | 29/825, 3.52% (2.46-5.00) | 308/4299, 7.16% (6.43-7.97) | OR 0.51 (95% CI: 0.30-0.84)<br>p<0.001 | 3 months |
| Kaminski et al. | 150/150mg (100%) | 41/333, 12.31% (9.21-16.28) | 42/97, 43.30% (33.88-53.23) | HR 0.011 (95% CI 0.063-0.198)<br>Log-rank: P <0.001 | Around 4-5 months |
| Chen et al. Preprint | 150/150mg (some),<br>300/300mg (some) | 102/1295, 7.88% (6.53-9.47) | 121/1295, 9.34% (7.88-11.05) |  | Between 1-7 months |
| Bruel et al. | Not provided | 4/29, 13.79% (5.50-30.56) | No control group |  | 23 days |
| Nguyen et al. | 150/150mg (100%) | 49/1112, 4.41% (3.35-5.78) | No control group |  | Median of 63 days (IQR: 49–73) |
| Benotmane et al. | 150/150mg (100%) | 7/63, 11.11% (5.49-21.20) | Infection rates not provided for control groups |  | Median of 29 days (IQR: 29-33) |
| Stuver et al. | 1x150/150mg (57.7%),<br>2x150/150mg (32.7%),<br>1x300/300mg (9.6%) | 2/52, 3.85% (1.06-12.98) | No control group |  | Median of 79 days |
| Benotmane et al. (2) | 150/150mg (100%) | 39/416, 9.38% (6.93-12.56) | No control group |  | 30 days |
| Ordaya et al. | 150/150mg (100%) | 8/674, 1.19% (0.60- | No control group |  | Median of 99 days (Range: 66- |
|  |  | 2.32) |  |  | 108) |
| **Al-Obaidi et al.** | 300/300mg (100%) | 6/98, 6.12% (2.84-12.72) | No control group |  | Median of 48 days (IQR: 27.5-69) |
| **Aqeel et al.** | 300/300mg (95%), 150/150mg (5%) | 4/21, 19.05% (7.67-40.00) | No control group |  | Mean of 98.6 ± 36.5 days |
| **Ocon et al.** | 300/300mg (88%), 150/150mg (12%) | 1/43, 2.33% (0.41-12.06) | No control group |  | Mean of 100 ± 33 days |
| **Calabrese et al.** | 150/150mg (some), 300/300mg (some) | 12/412, 2.91% (1.67-5.02) | No control group |  | Range of 13-84 days |
| **Woopen et al.** | 2x150/150mg (100%) | 0/6, 0.00% (0.00-39.03) | No control group |  | About 2 months |

### Study quality

The GRADE tool was used to assess the certainty of the evidence for the primary outcome in each of the nine included studies. Supplementary Table 1 provide a summary of the GRADE scores of each study, and the reasons for each negative scoring.

The PROVENT study, the only included randomised controlled trial, was assessed according to the Cochrane risk of bias assessment and deemed to have a low risk of bias (full summary in Appendix 2)^8,32^.

### Clinical effectiveness against breakthrough COVID-19 infections

Tixagevimab/Cilgavimab is a combination of two human monoclonal antibodies targeted against the surface spike protein of SARS-CoV-2. The antibodies have a modification to their Fc receptor to ensure a longer duration of action and enable use as a prophylactic antibody therapy^6^. As such, the primary clinical effectiveness measure is the prevention of breakthrough COVID-19 infections. We identified 17 studies that reported on this measure.

The original phase 3 licensing study was the PROVENT study. This was a randomised controlled study, randomising 5197 patients between Tixagevimab/Cilgavimab or placebo. This demonstrated a relative risk reduction of 76.7% (95% confidence interval [CI], 46.0-90.0; p<0.001) after a median of 83 days from Tixagevimab/Cilgavimab administration (150/150mg) compared to placebo. The secondary outcome findings after a median of 196 days revealed a symptomatic COVID-19 infection risk reduction of 82.8% (CI: 65.8-91.4%) ^8^.

Subsequent to the phase 3 study, sixteen real-world post-licensing studies have been performed assessing clinical effectiveness against breakthrough infections. Five were performed in reference to a control group^16–20^, and reported clinical outcomes from propensity-matched U.S. army veterens^16^, solid organ transplant recipients^17^, kidney transplant recipients^19^ and from generally immunocompromised patients^18,20^. Four studies showed a reduction in COVID-19 infections in the Tixagevimab/Cilgavimab group compared to control groups ^16– 19^. Chen et al compared the same participant population before and after Tixagevimab/Cilgavimab, finding a reduced infection rate post intervention^20^.

It is important to note that three studies^8,18,19^ included 150mg/150mg Tixagevimab/Cilgavimab dosages, while three studies also included participants who received 300/300mg dosages^16,17^. Al Jurdi et al. also included a 450/450mg Tixagevimab/Cilgavimab recipient^17^. Al Jurdi et al. found that, during a mean follow-up of 87 ±30 days, the rate of COVID-19 infection breakthrough was higher amongst those who received the 150/150mg Tixagevimab/Cilgavimab dose compared to the 300/300mg dose (p=0.025)^17^.

Of the eleven cohort studies without controls, six were prospective^26–31^ and five were retrospective^12,22–25^. Infection rates between 0-19% over a 0.5-4-month period were reported in these cohort studies ^12,22–31^. Nguyen et al. reported a low infection rate of 4.4% (49/1112) after a median follow-up of 63 days. Of the 29 cases with viral sequencing from Nguyen’s paper, all the cases were found to be Omicron infections. At the time of the study, the mean weekly incidence rate in Ile-de-France was 1,669 infections in 100,000 inhabitants, whereas amongst the study population the incidence rate was 530 infections in 100,000 inhabitants^27^. Breakthrough infection rates reported across each study are summarised in Table 1.

Overall, the clinical effectiveness of COVID-19 breakthrough infections in patients who received group Tixagevimab/Cilgavimab compared to a control group was 40.47% (CI 29.82-49.67) (p<0.0001).

### Clinical effectiveness against COVID-19 hospitalisation

Tixagevimab/Cilgavimab is believed to reduce the severe sequelae of hospitalisation in a similar fashion to coronavirus vaccines^15,33^. In the original PROVENT trial, one patient in the control group was hospitalised with a severe infection (needing non-invasive ventilation or high-flow oxygen) and four patients in the control group were hospitalised with a severe infection. There were no COVID-19 hospitalisations in the Tixagevimab/Cilgavimab group^8^.

Of the seventeen studies included in this review, sixteen have reported on hospitalisation of patients who received Tixagevimab/Cilgavimab. Six of these studies included corresponding controls^8,16–20^. All of these studies demonstrated reduced rates of hospitalisation in patients receiving Tixagevimab/Cilgavimab. The rates of hospitalisations in the ten cohort studies ranged between 0-9% over a 0.5-4-month period^12,22–25,27–31^. Hospitalisation rates reported across each study are summarised in Table 2.

**Table 2:** Secondary outcome hospitalisations summary ^8,12,16–20,22–25,27–31^. (RRR: relative risk ratio; HR: hazard ratio; IRR: incidence risk ratio; OR: odds ratio; IQR: interquartile range)

| Paper | Hospital admission<br>Hospitalisations/ no of participants<br>% (95% CI) |  | Relative effect (95% CI) or significance | Time course |
| --- | --- | --- | --- | --- |
|  | Intervention | Control |  |  |
| **Levin et al.** | 0/3441, 0.00% (0.00-0.11) | 3/1731, 0.17% (0.06-0.51) |  | Median of 83 days |
|  | 0/3441, 0.00% (0.00-0.11) | 7/1731, 0.40% (0.20-0.83) |  | Median of 196 days |
| **Young-Xu et al. (preprint)** | <9/1733, 0.52% (0.27-0.98) | 38/6354, 0.60% (0.44-0.82) | HR 0.13; 95%CI, 0.02-0.99<br>IRR: 0.10 (0.05-0.22) | Over 4 months |
| **Al Jurdi et al.** | 1/222, 0.45% (0.08-2.51) | 6/222, 2.70% (1.24-5.77) |  | Mean of 87 ± 30 days after Tixagevimab/Cilgavimab; mean of 82 ± 28 after control |
| **Kertes et al.** | 1/825, 0.12% (0.02-0.68) | 27/4299, 0.63% (0.43-0.91) | p=0.07 | 3 months |
| **Kaminski et al. Hospitalisation** | 4/333, 1.20% (0.47-3.05) | 11/97, 11.34% (6.45-19.17) | HR, 0.046; 95% CI, 0.013–0.158; P < 0.001 | Around 4-5 months |
| **Kaminski et al. Intensive care** | 2/333, 0.60% (0.16-2.16) | 6/97, 6.19% (2.87-12.84) | HR, 0.045; 95% CI, 0.008–0.240; P < 0.001 | Around 4.5 months |
| **Chen et al. (preprint)** | 6/1295, 0.46% (0.21-1.01) | 36/1295, 2.78% (2.01-3.82) | rate ratio (RR) = 0.20; 95% confidence | Between 1-7 months |
| Hospitalisation |  |  | intervals (CI) = 0.09-0.45; p <0.001 |  |
| **Chen et al. (preprint)**<br>Intensive care | 0/1295,<br>0.00% (0.00-0.30) | 8/1295,<br>0.62% (0.31-1.21) |  | Between 1-7 months |
| **Nguyen et al.** | 6/1112,<br>0.54% (0.25-1.17) | No control group |  | Median of 63 days (IQR: 49-73) |
| **Benotmane et al. (preprint)** | 2/63,<br>3.17% (0.87-10.86) | Not provided for control group |  | Median of 29 days (IQR: 29-33) |
| **Stuver et al.** | 0/52,<br>0.00% (0.00-6.88) | No control group |  | Median of 79 days |
| **Benotmane et al. (2)**<br>Hospitalisation | 14/416,<br>3.37% (2.02-5.57) | No control group |  | 30 days |
| **Benotmane et al. (2)**<br>Intensive care | 3/416,<br>0.72% (0.25-2.10) | No control group |  | 30 days |
| **Ordaya et al.** | 2/674,<br>0.30% (0.08-1.08) | No control group |  | Median of 99 days (Range: 66-108) |
| **Al-Obaidi et al.** | 42/463,<br>9.07% (6.78-12.03) | No control group |  | Median of 48 days (IQR: 27.5-69) |
| **Aqeel et al.** | 0/21,<br>0.00% (0.00-15.46) | No control group |  | Mean of 98.6 ± 36.5 days |
| **Ocon et al.** | 0/43,<br>0.00% (0.00-8.20) | No control group |  | Mean of 100 ± 33 days |
| **Calabrese et al.** | 1/412,<br>0.24% (0.04-1.36) | No control group |  | Range of 13-84 days |
| **Woopen et al.** | 0/6,<br>0.00% (0.00-39.03) | No control group |  | About 2 months |

Overall, the clinical effectiveness of Tixagevimab/Cilgavimab against COVID-19 hospital admissions was 69.23% (CI: 50.78-81.64) (p<0.00001).

Three papers also reported numbers of intensive care hospitalisation^12,19,20^, two of which were comparative studies^19,20^. Kaminski et al. reported a statistically significant reduced number in the Tixagevimab/Cilgavimab intervention group of 0.6% (2/333 cases) compared to 6.2% (6/97) in the control group^19^. Chen et al. reported a decreased rate of intensive care hospitalisations after Tixagevimab/Cilgavimab administration (0.6% before, 0.0% afterwards)^20^. The clinical effectiveness against COVID-19 intensive care admission from these two studies was 87.89% (CI: 47.12-98.66) (p=0.0008).

### Clinical effectiveness in preventing COVID-19 mortality

Prior to this systematic review, there was limited evidence about the clinical effectiveness of Tixagevimab/Cilgavimab in preventing COVID-19 mortality.

In the original PROVENT study, there were no coronavirus deaths in the patients treated with Tixagevimab/Cilgavimab, and two deaths in the control group (2/3461, 0.06%)^8^.

Of the real-world evaluation studies reviewed, sixteen reported on mortality in patients who received Tixagevimab/Cilgavimab, with six including controls ^8,16–20^. Of these, in five studies, the rates of both COVID-19-specific deaths^8,17,19^, and all-cause mortality^8,16,18^ were lower in the Tixagevimab/Cilgavimab-treated groups compared to controls. Chen et al. found no difference in COVID-19-related mortality before and after Tixagevimab/Cilgavimab^20^. In the ten cohort studies without controls, rates of COVID-19-specific deaths were very low, ranging between 0.00-0.48% over a 0.5-4-month period^12,22–25,27–31^. Eight of these studies reported mortality rates of 0.00% ^22–25,28–31^. Al-Obaidi et al reported an all-cause mortality rate of 0.86% after a median of 53 days and was the only non-comparative cohort study which included all-cause mortality^23^. Mortality rates reported across each study are summarised in Table 3.

**Table 3:** Secondary outcome mortality summary ^8,12,16–20,22–25,27–31^. (RRR: relative risk ratio; HR: hazard ratio; IRR: incidence risk ratio; OR: odds ratio; IQR: interquartile range)

| Paper | Mortality type | Mortality<br>Total mortality/ no of participants<br>% (95% CI) |  | Relative effect (95% CI) or significance | Time course |
| --- | --- | --- | --- | --- | --- |
|  |  | Intervention (95% CI) | Control |  |  |
| **Levin et al.** | All-cause | 9/1341,<br>0.67% (0.35-1.27) | 7/1731,<br>0.40% (0.20-0.83) |  | Median of 196 days |
|  |  | 4/3461,<br>0.12% (0.04-0.30) | 4/1736,<br>0.23% (0.09-0.59) |  | Median of 83 days |
| **Levin et al.** | Covid-19-related | 0/3461,<br>0.00% (0.00-0.11) | 2/3461,<br>0.06% (0.02-0.21) |  |  |
| **Young-Xu et al. (preprint)** | All-cause | <9/1733,<br>0.52% (0.27-0.98) | 99/6354,<br>1.56% (1.28-1.89) | HR 0.36;<br>95%CI, 0.18-0.73 | Over 4 months |
| **Al Jurdi et al.** | Covid-19-related | 0/222,<br>0.00% (0.00-1.70) | 3/222,<br>1.35% (0.46-3.90) |  | Mean of 87 ± 30 days after<br>Tixagevimab/Cilgavimab;<br>mean of 82 ± 28 after control |
| **Kertes et al.** | All-cause | 0/825,<br>0.00% (0.00-0.46) | 40/4299,<br>0.93% (0.68-1.26) | p=0.005 | 3 months |
| **Kaminski et al.** | Covid-19-related | 1/333,<br>0.30% (0.05-1.68) | 2/97,<br>2.06% (0.57-7.21) | HR, 0.076;<br>95% CI,<br>0.005–1.161;<br>p = 0.066 | Around 4-5 months |
| **Chen et al. (preprint)** | Covid-19-related | 0/1295,<br>0.00% (0.00-0.30) | 0/1295,<br>0.00% (0.00-0.30) |  | Between 1-7 months |
| **Nguyen et al.** | Covid-19-related | 2/1112,<br>0.18% (0.05-0.65) | No control group |  | Median of 63 days<br>(IQR: 49–73) |
| **Benotmane et al. (preprint)** | Covid-19-related | 0/63,<br>0.00% (0.00-5.75) | Mortality not provided for control group |  | Median of 29 days<br>(IQR: 29-33) |
| **Stuver et al.** | Covid-19-related | 0/52,<br>0.00% (0.00-6.88) | No control group |  | Median of 79 days |
| **Benotmane et al. (2)** | Covid-19-related | 2/416,<br>0.48% (0.13-1.74) | No control group |  | 30 days |
| **Ordaya et al.** | Covid-19-related | 0/674,<br>0.00% (0.00-0.57) | No control group |  | Median of 99 days<br>(Range: 66-108) |
| **Al-Obaidi et al.** | All-cause | 4/463,<br>0.86% (0.34-2.20) | No control group |  | Median of 53 days<br>(IQR 18-91) |
| **Al-Obaidi et al.** | Covid-19-related | 0/463,<br>0.00% (0.00-0.82) | No control group |  | Median of 53 days<br>(IQR 18-91) |
| **Aqeel et al.** | Covid-19-related | 0/21,<br>0.00% (0.00-15.46) | No control group |  | Mean of 98.6 ± 36.5 days |
| **Ocon et al.** | Covid-19-related | 0/43,<br>0.00% (0.00-8.20) | No control group |  | Mean of 100 ± 33 days |
| **Calabrese et al.** | Covid-19-related | 0/412,<br>0.00% (0.00-0.92) | No control group |  | Range of 13-84 days |
| **Woopen et al.** | Covid-19-related | 0/6,<br>0.00% (0.00-39.03) | No control group |  | About 2 months |

The overall clinical effectiveness percentages of Tixagevimab/Cilgavimab for patients in studies reporting all-cause mortality ^8,16,18^ and COVID-19-specific mortality ^8,17,19,20^ were 81.29% (66.93-90.28) p<0.0001 and 86.36% (CI: 6.21-99.70) (p=0.0351), respectively.

### Neutralising ability of patient sera

Five included studies analysed the neutralising ability of patient sera after Tixagevimab/Cilgavimab administration^8,12,26,28,29^. The phase III PROVENT trial showed that the neutralising antibody titres to the wild-type SARS-CoV-2 receptor-binding domain (RBD) after Tixagevimab/Cilgavimab was significantly higher than the convalescent plasma antibody titres of healthy patients from the phase I trial^8^.

However, analysis from different papers shows varying amounts of neutralisation of different SARS-CoV-2 variants. Bruel et al. found that, 15-30 days post Tixagevimab/Cilgavimab (dosage not provided), sera from 8 of the 11 participants had neutralising activity against Omicron variant BA.1, and all 11 were able to neutralise BA.2. Neutralisation of SARS-CoV-2 delta variant was nine-fold higher than BA.2 neutralisation in the 11 Tixagevimab/Cilgavimab-treated participants^26^. After a median of 33 and 29 days of 150/150mg Tixagevimab/Cilgavimab, respectively, both Stuver’s and Benotmane’s studies showed minimal Omicron-RBD neutralisation^28,29^. Stuver found that, although participant sera had high anti-spike IgG levels and effective wildtype-RBD neutralisation, the median Omicron-RBD neutralisation failed to reach the positive cut-off value^29^. Similarly, Benotmane found that only 9.5% of patient plasma (6/63) was able to neutralise Omicron variant BA.1 RBD, compared to 71% of patients who were infected naturally with SARS-CoV-2^28^. Benotmane et al. (2) also found that, of the five kidney transplant recipients with breakthrough COVID-19 infection tested, all 5 had minimal anti-RBD IgG levels against SARS-CoV-2 BA.1 (<3500BAU/mL)^12^.

Of note, Stuver’s study showed that those who were administered two 150/150mg or a 300/300mg Tixagevimab/Cilgavimab dose had significantly higher Omicron-RBD neutralisation (p=0.003), with 90% of patient sera neutralisation levels above the positive cut-off value^29^.

## Discussion

This systematic review has assessed all clinical studies reporting on the clinical effectiveness of Tixagevimab/Cilgavimab. Prophylactic therapy showed clinical effectiveness against coronavirus breakthrough infections of 40.47%, COVID-19 hospitalisation of 69.23%, ITU admission of 87.89%, all-cause mortality of 81.29% and COVID-19-specifc mortality of 86.36%.

Globally, countries are still grappling with ongoing waves of COVID-19, often driven by new coronavirus variants. Population-scale coronavirus vaccination programmes have been effective in preventing the most severe sequelae in the majority of healthy individuals. However, these have not been able to completely prevent transmission amongst individuals, many of whom may have minor or asymptomatic infections^34^.

It is also evident that there are groups of patients who remain at increased risk of severe outcomes, including immunocompromised individuals. While booster vaccinations have been shown to provide moderate protection for some of these patients^35^, many are unable to elicit appropriate immune responses following multiple coronavirus boosters. Thus, booster vaccination alone is unlikely to equitably mitigate the excess risk in these patient groups.

The long-acting prophylactic antibody therapy Tixagevimab/Cilgavimab has been shown to be an effective measure for immunocompromised patient groups. The original licensing study, PROVENT has changed clinical practice globally, and 32 countries now use prophylactic antibody therapy in conjunction with vaccination as their pandemic standard of care^36^. However, some uncertainty does remain. The original PROVENT study was performed prior to the recent SARS-COV-2 waves driven by the omicron variants. Furthermore, when new SARS-CoV-2 variants emerge, there are invariably calls for new randomised clinical trials. This systematic review is therefore important and is the first to review the existing body of evidence as to the clinical effectiveness of this treatment during the omicron era.

This systematic review included 17 large scale human studies, including a total of 24,773 immunocompromised participants, of whom 10,775 received Tixagevimab/Cilgavimab as a prophylactic antibody therapy. While our findings are generally positive, it should be acknowledged that the real-world studies differed in quality. Many studies did not have perfect controls and comprised different patient groups. It is also the case that the studies report on current real-world effectiveness against current SARS-CoV-2 variants and cannot forecast clinical effectiveness against future SARS-CoV-2 variants.

From a wider perspective, we must note that surrogate measures of clinical effectiveness such as laboratory neutralisation assays using synthetic or derived virus have illustrated potential resistance to Tixagevimab/Cilgavimab^37,38^. Indeed, antibody therapies have additional functions other than simple neutralisation, including regulation of cell-mediated cytotoxicity, agglutination and opsonisation. The results of this systematic review suggest that the higher 300mg/300mg dosage of Tixagevimab/Cilgavimab, which is recommended by the manufacturer^11^, could reduce the chance of SARS-CoV-2 Omicron strain resistance. We believe surrogate markers of clinical effectiveness of clinical effectiveness must be validated prior to extrapolating clinical conclusions. Indeed, two of the studies included in this paper found a high interindividual variability in neutralising antibody levels post-Tixagevimab/Cilgavimab ^24,28^, with Aqeel et al. reporting that change in IgG levels post-Tixagevimab/Cilgavimab was only reported in one patient^24^.

In summary, this systematic review has illustrated that there is now a growing body of real-world evidence validating the original phase 3 study as to the clinical effectiveness of Tixagevimab/Cilgavimab and demonstrating effectiveness in the Omicron era. It is critically important that larger-scale and better-controlled pilots and evaluations are performed to highlight the significant clinical benefit of prophylactic antibody treatment in immunocompromised groups. Further co-ordinated work is required to ensure that the risk to these vulnerable individuals from coronavirus is further mitigated and there is an ongoing pipeline of long-acting medical products which could provide even higher levels of protection as the pandemic continues.

## Data Availability

All data produced in the present work are contained in the manuscript and/or cited publications.

---

**Supplementary Table 1:** primary outcome GRADE scores^8,12,16–20,22–31^.

| Paper | Study population | GRADE score = Level<br>[Reason key] |
| --- | --- | --- |
| Levin et al. | Adults at increased risk of SARS-CoV-2 infection | (+)(+)(+)(-) = Level 3<br>[a] |
| Young-Xu et al. (preprint) | Immuno-compromised U.S. Veterans | (+)(+)(-)(-) = Level 2<br>[b] |
| Al Jurdi et al. | Solid organ transplant recipients | (+)(+)(-)(-) = Level 2<br>[b] |
| Kertes et al. | Immunocompromised patients at Maccabi HealthCare Services, Israel | (+)(+)(-)(-) = Level 2<br>[b] |
| Kaminski et al. | Kidney transplant recipients | (+)(+)(-)(-) = Level 2<br>[b] |
| Chen et al. (preprint) | Immunocompromised patients | (+)(+)(-)(-) = Level 2<br>[b] |
| Bruel et al. | Immunocompromised individuals. 18/29 participants had previously been administered Ronapreve, a mean of 35 days before Tixagevimab/Cilgavimab administration | (+)(-)(-)(-) = Level 1<br>[b, c] |
| Nguyen et al. | Immunocompromised patients | (+)(-)(-)(-) = Level 1<br>[b, d] |
| Benotmane et al. (preprint) | Kidney transplant recipients | (-)(-)(-)(-) = Level 0<br>[b, c, d] |
| Stuver et al. | Patients with haematological malignancies | (+)(-)(-)(-) = Level 1<br>[b, d] |
| Benotmane et al. (2) | Kidney transplant recipients | (+)(-)(-)(-) = Level 1<br>[b, d] |
| Ordaya et al. | Immunocompromised patients | (+)(-)(-)(-) = Level 1<br>[b, d] |
| Al-Obaidi et al. | Immunocompromised patients | (-)(-)(-)(-) = Level 0<br>[a, b, d] |
| Aqeel et al. | Rituximab-treated antineutrophil cytoplasmic antibody vasculitis patients | (+)(-)(-)(-) = Level 1<br>[b, d] |
| Ocon et al. | Rheumatological patients on Rituximab | (+)(-)(-)(-) = Level 1<br>[b, d] |
| Calabrese et al. | Those with immune-mediated inflammatory disease, undergoing B cell depleting therapy, and those with inborn errors of humoral immunity | (+)(-)(-)(-) = Level 1<br>[b, d] |
| Woopen et al. | Multiple sclerosis patients | (-)(-)(-)(-) Level 0<br>[b, c, d] |

## Appendix 1

### Risk of bias assessment: Levin et al.^8,32^

1. Risk of bias arising from the randomization process
  - Was the allocation sequence random? Yes, via a random number generator
  - Was the allocation sequence concealed until participants were enrolled and assigned to interventions? Yes
  - Did baseline differences between intervention groups suggest a problem with the randomization process? No baseline differences
    - **Low risk of bias**
2. Risk of bias due to deviations from the intended interventions (effect of assignment to intervention)
  - Were participants aware of their assigned intervention during the trial? Yes, for the consideration of vaccination status
  - Were carers and people delivering the interventions aware of participants assigned intervention during the trial? The pharmacists were unblinded, but the investigators and staff involved in follow-up and patient care were blinded
  - Were there deviations from the intended intervention that arose because of the trial context? No, the proportion of participants who were aware of their assignments were balanced between both groups.
    - **Low risk of bias**
3. Missing outcome data
  - Were data for this outcome available for all, or nearly all, participants randomized? No, some data missing-out of 5,197 participants, only 4,685 were evaluated. Reasons given include loss to follow-up or discontinuation, but further information is not provided
  - Is there evidence that the result was not biased by missing outcome data? Not clear
  - Could missingness in the outcome depend on its true value? Unlikely
    - **Unclear risk of bias**
4. Risk of bias in measurement of the outcome
  - Was the method of measuring the outcome inappropriate? No
  - Could measurement or ascertainment of the outcome have differed between intervention groups? Probably not
  - Were outcome assessors aware of the intervention received by study participants? No
    - **Low risk of bias**
5. Risk of bias in selection of the reported result
  - Were the data that produced this result analysed in accordance with a pre-specified analysis plan that was finalized before unblinded outcome data were available for analysis? Details for the primary analysis were outlined in the protocol, but this was not the case for the secondary, 6-month analysis.
  - Is the numerical result being assessed likely to have been selected, on the basis of the results, from multiple eligible outcome measurements (e.g., scales, definitions, time points) within the outcome domain? Unlikely
  - Is the numerical result being assessed likely to have been selected, on the basis of the results, from multiple eligible analyses of the data? No for the primary; ulcer for the secondary outcomes
    - **Unclear risk of bias**

## Appendix 2 Study characteristics

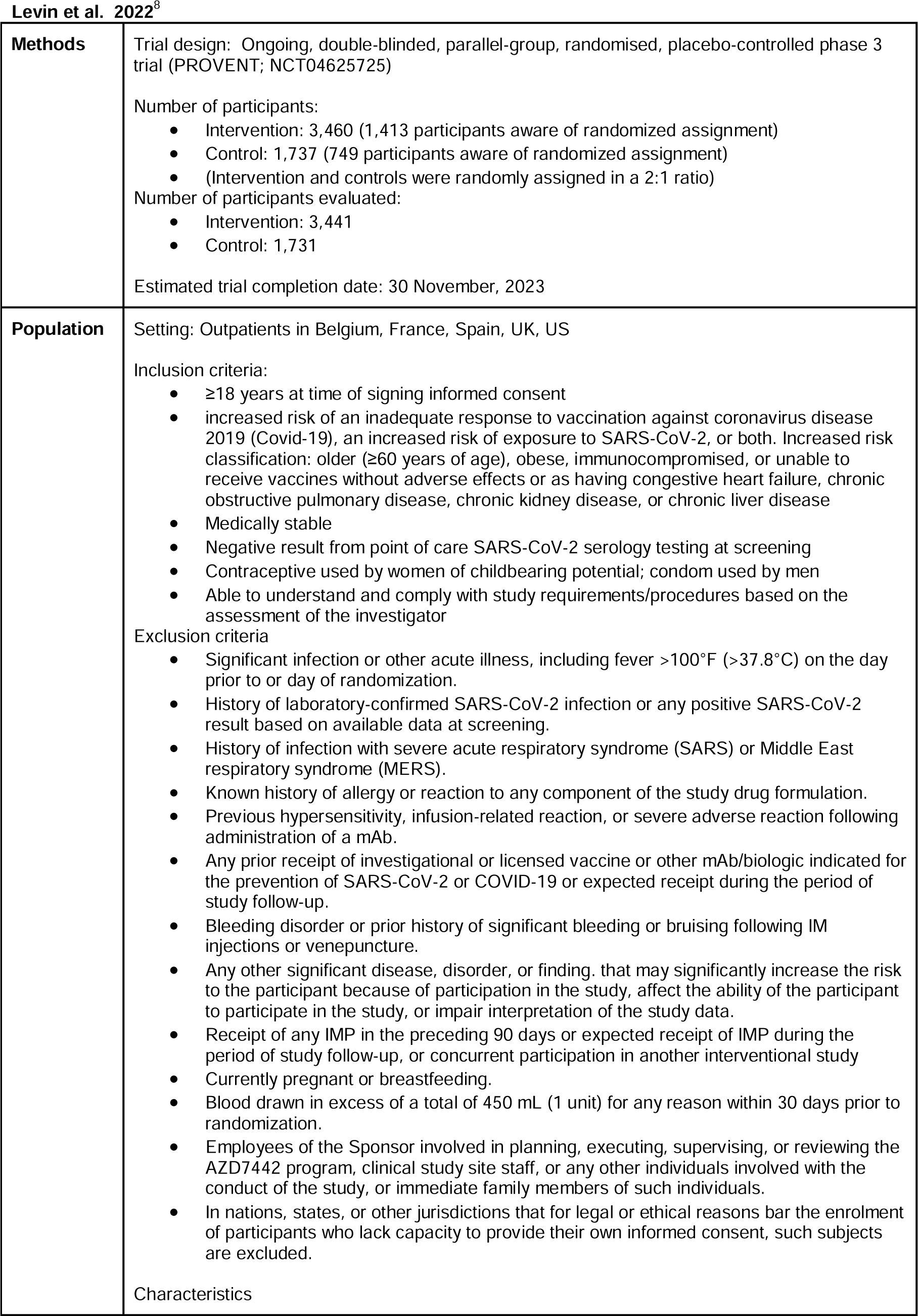
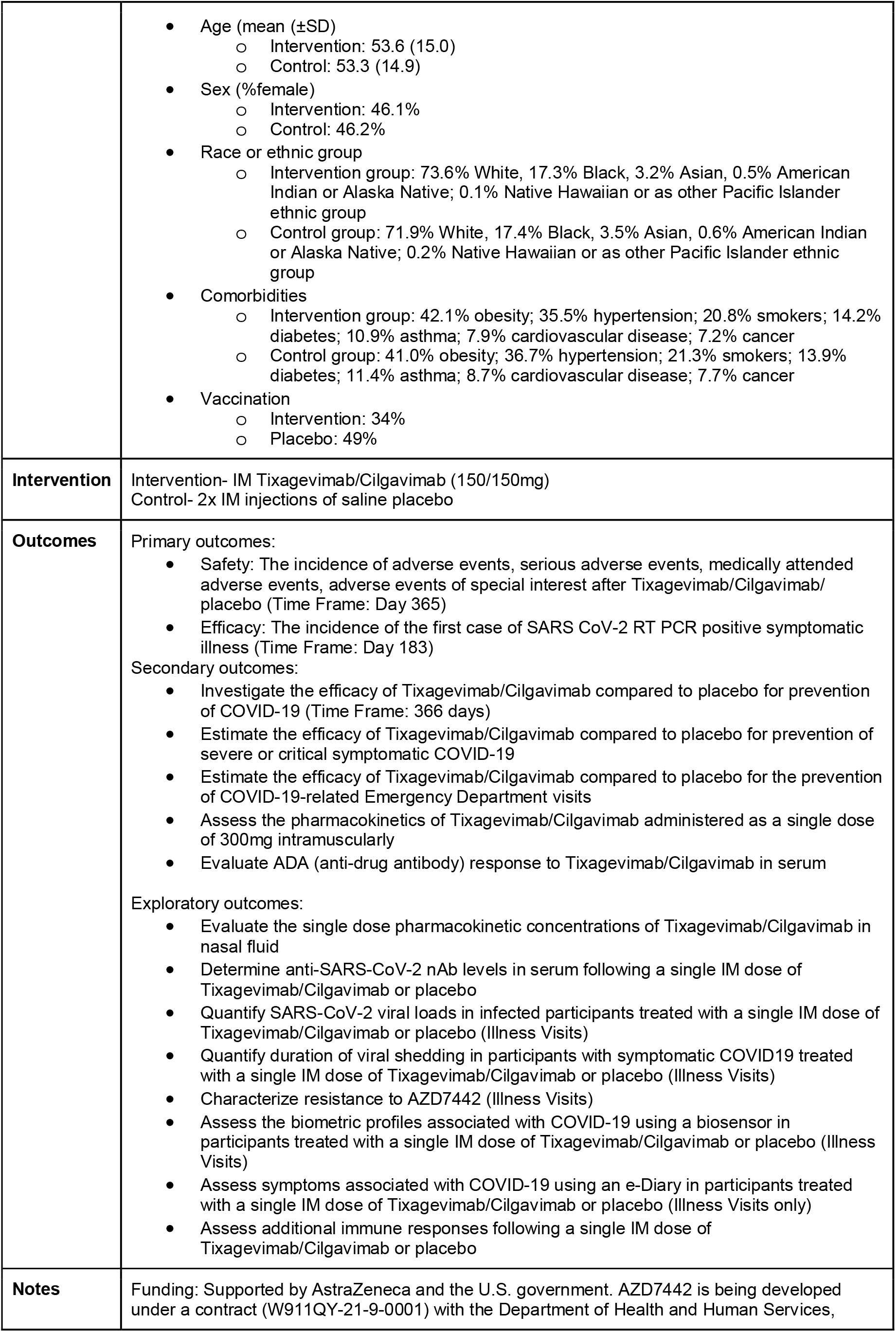
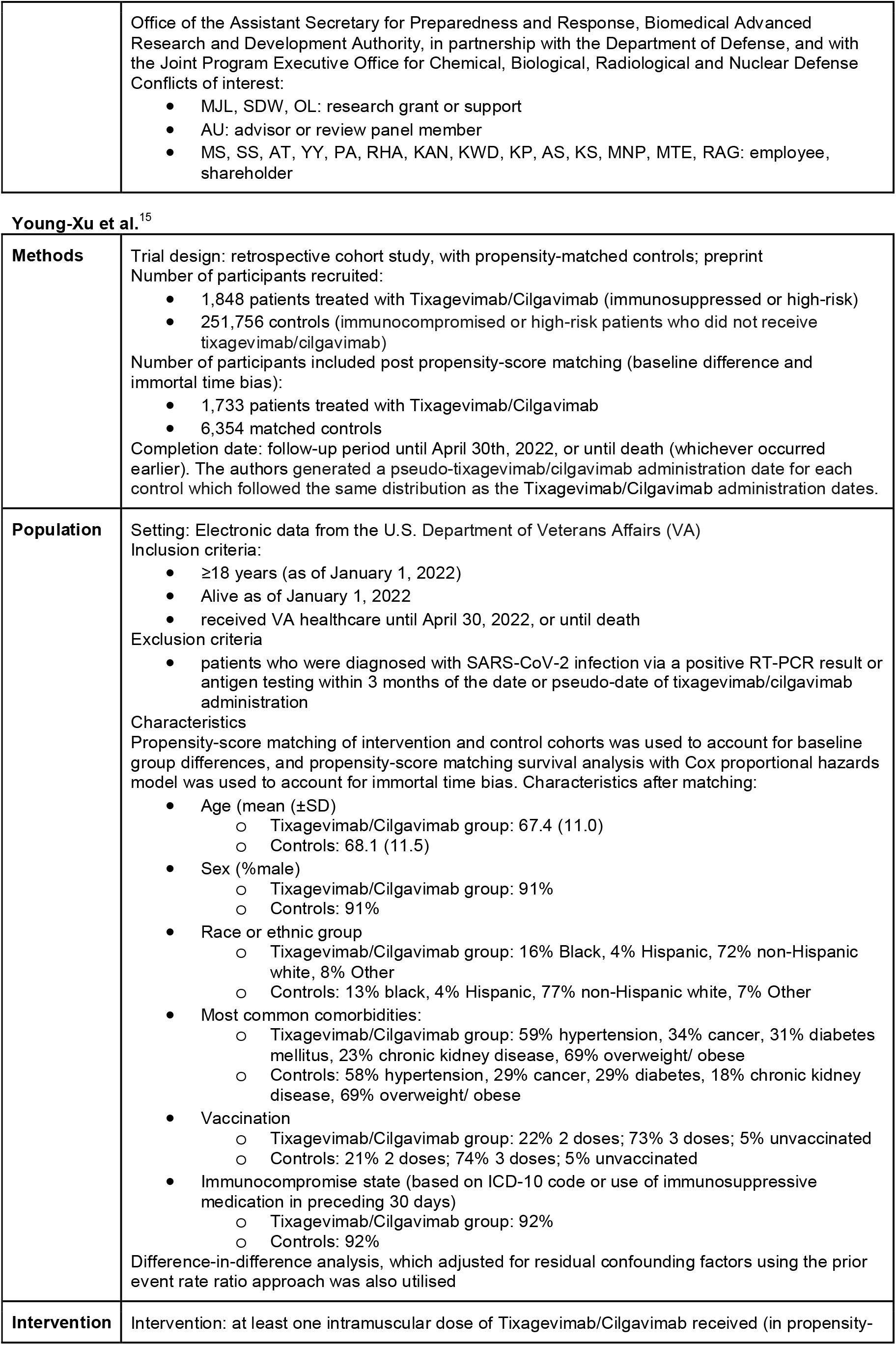
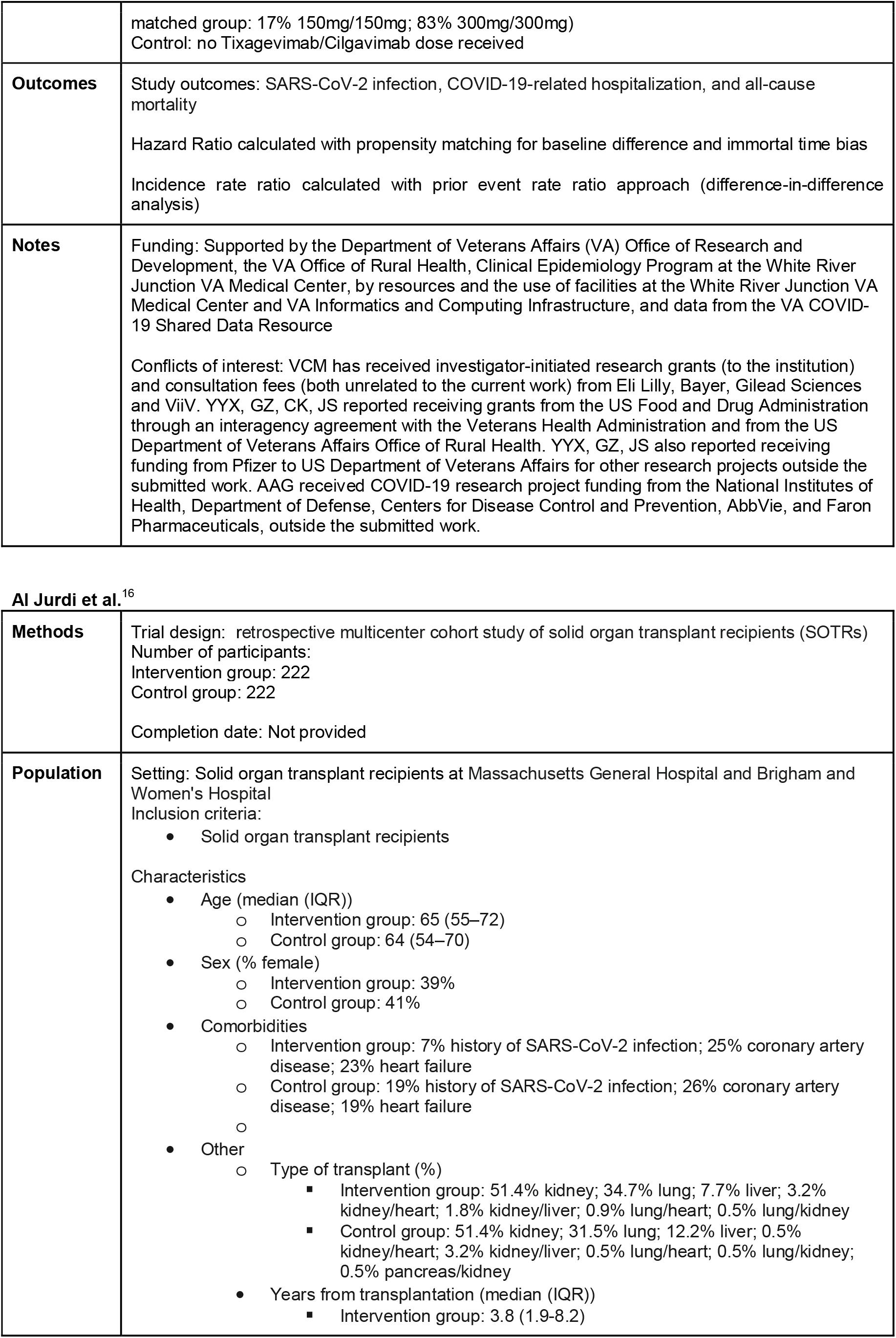
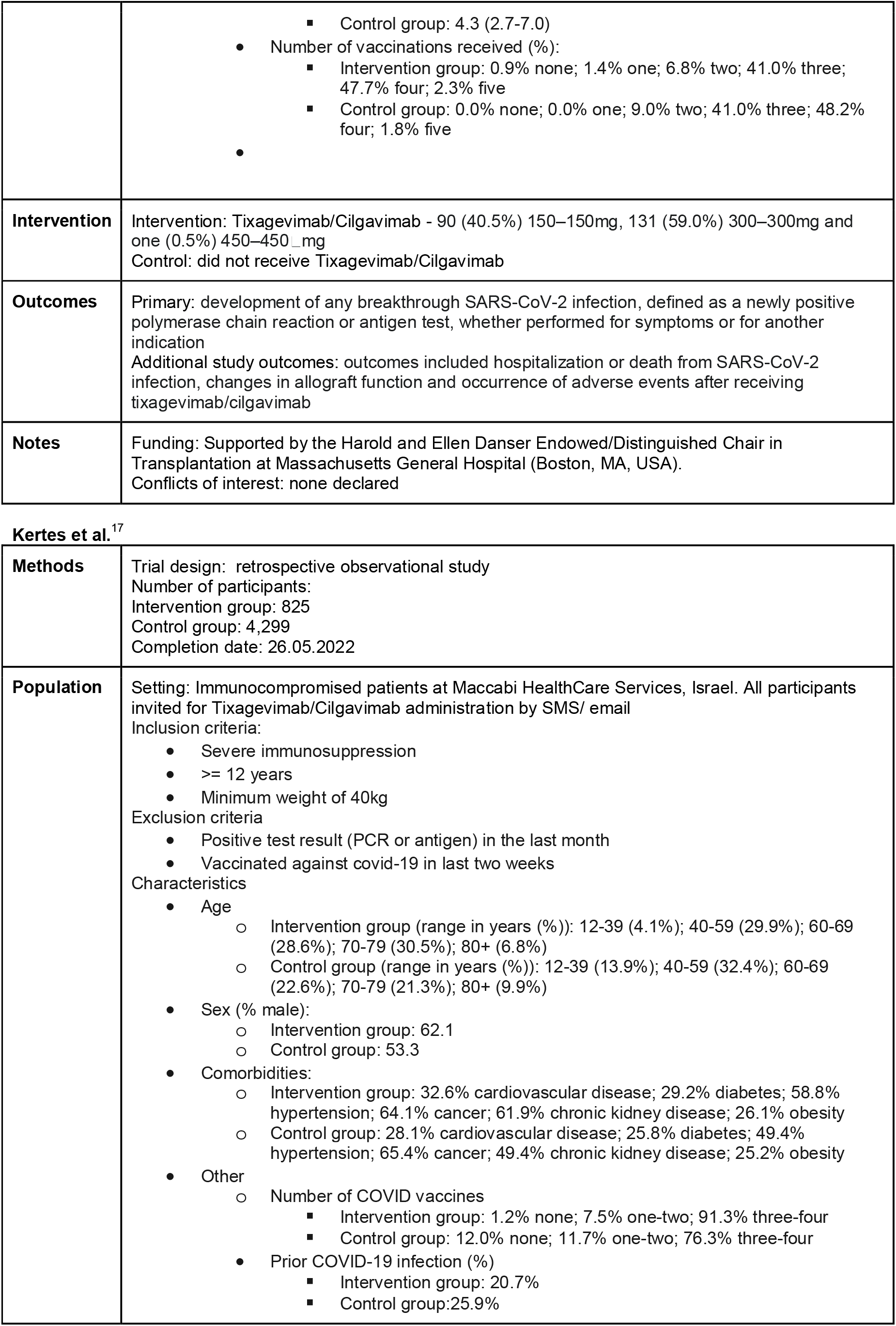
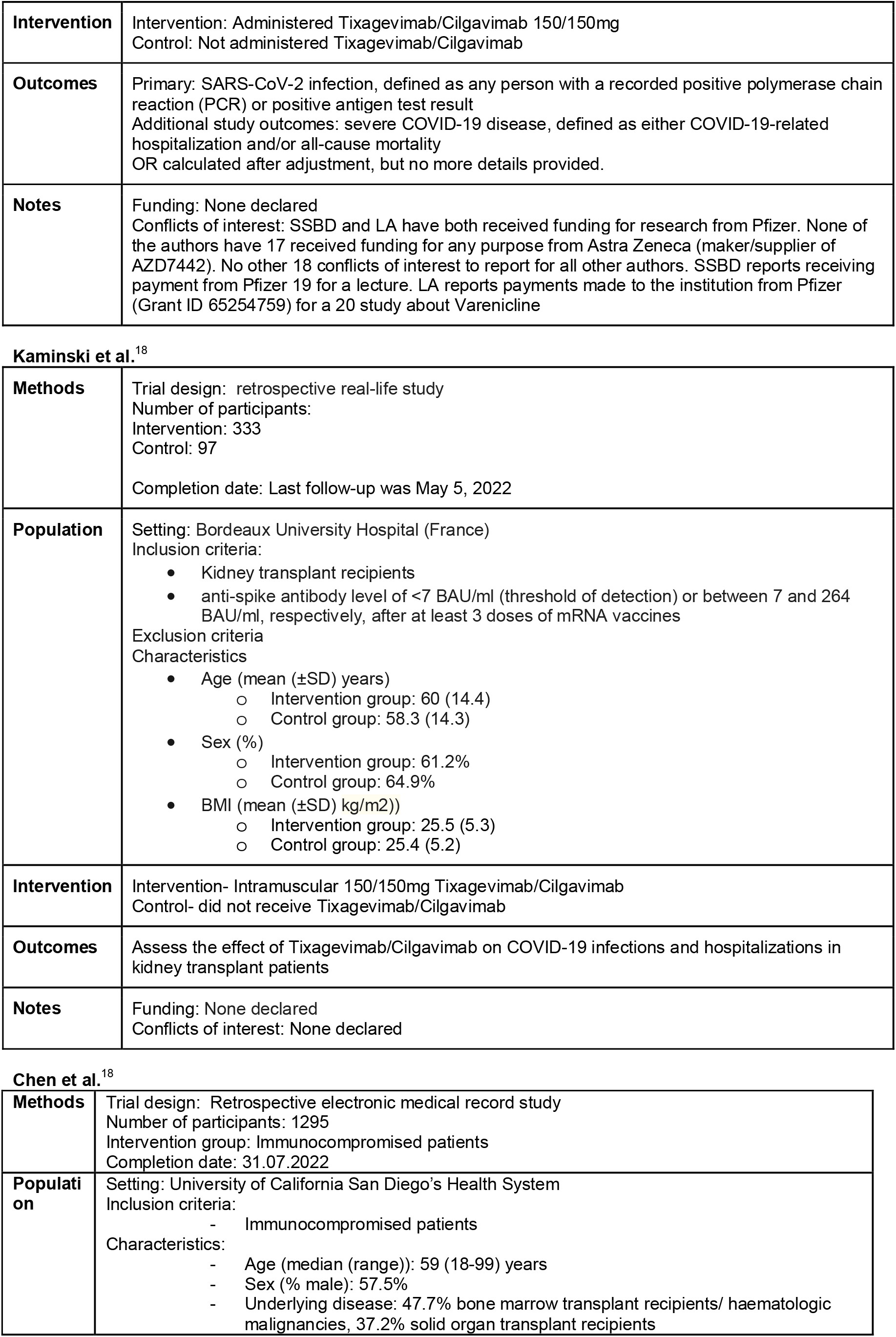
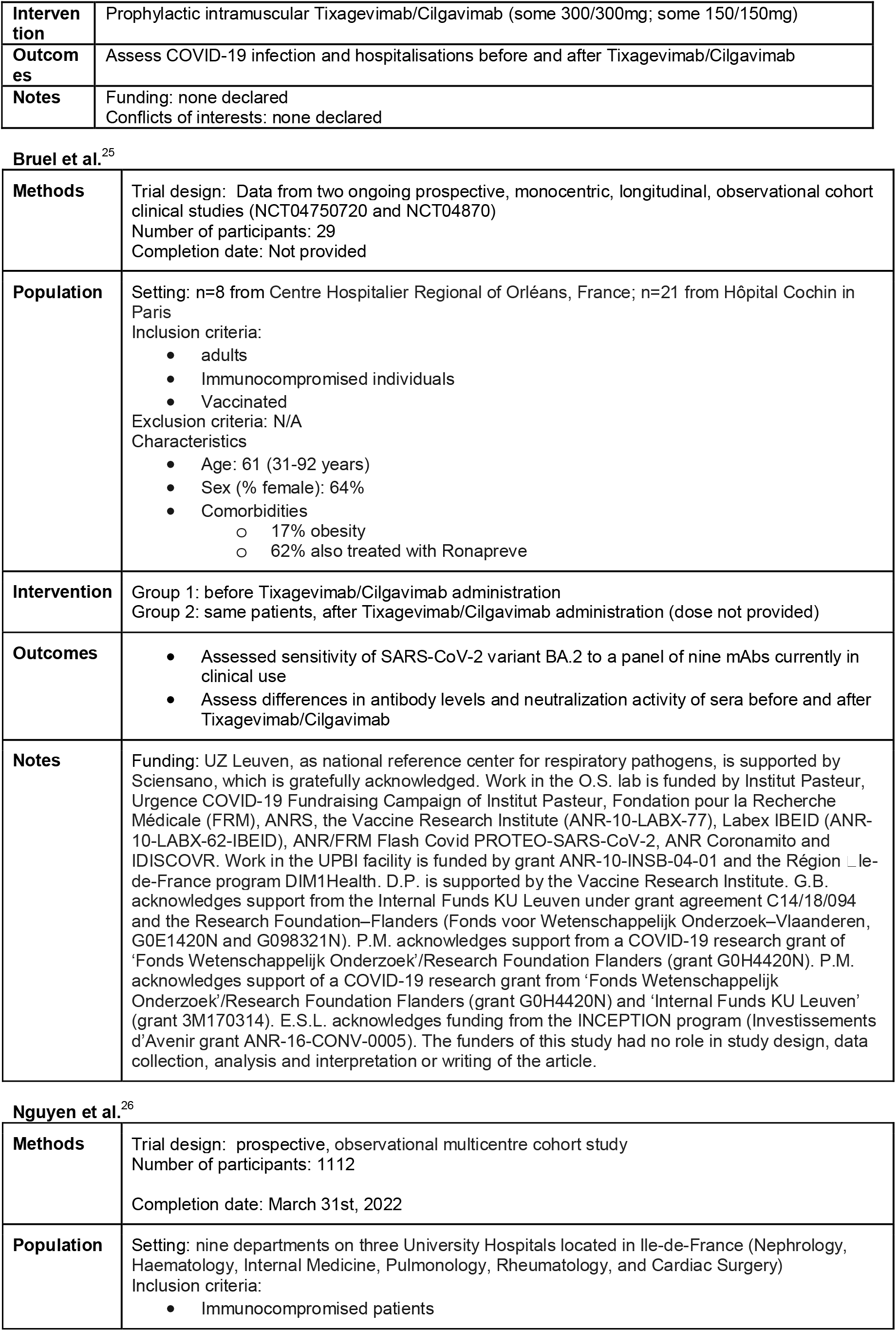
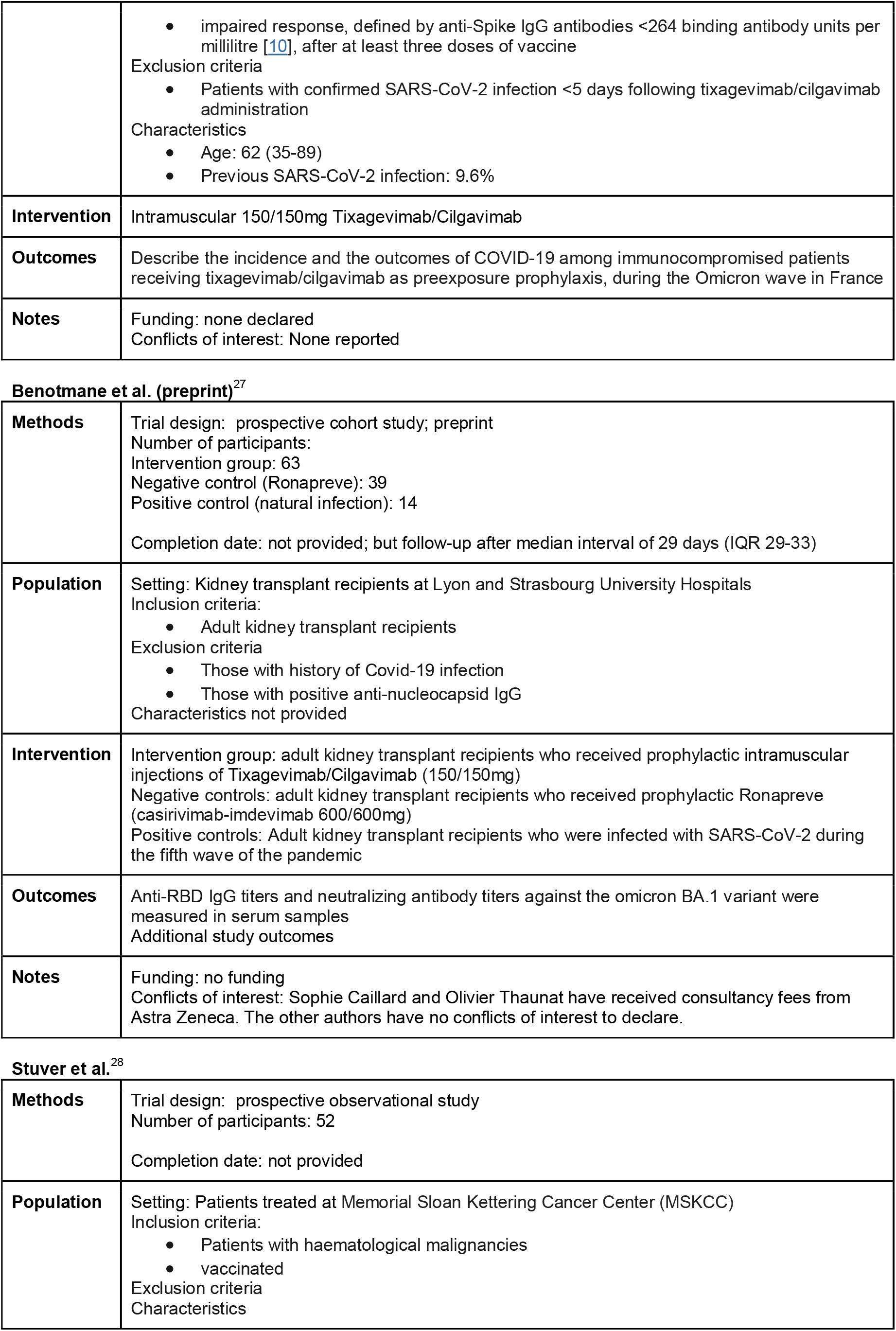
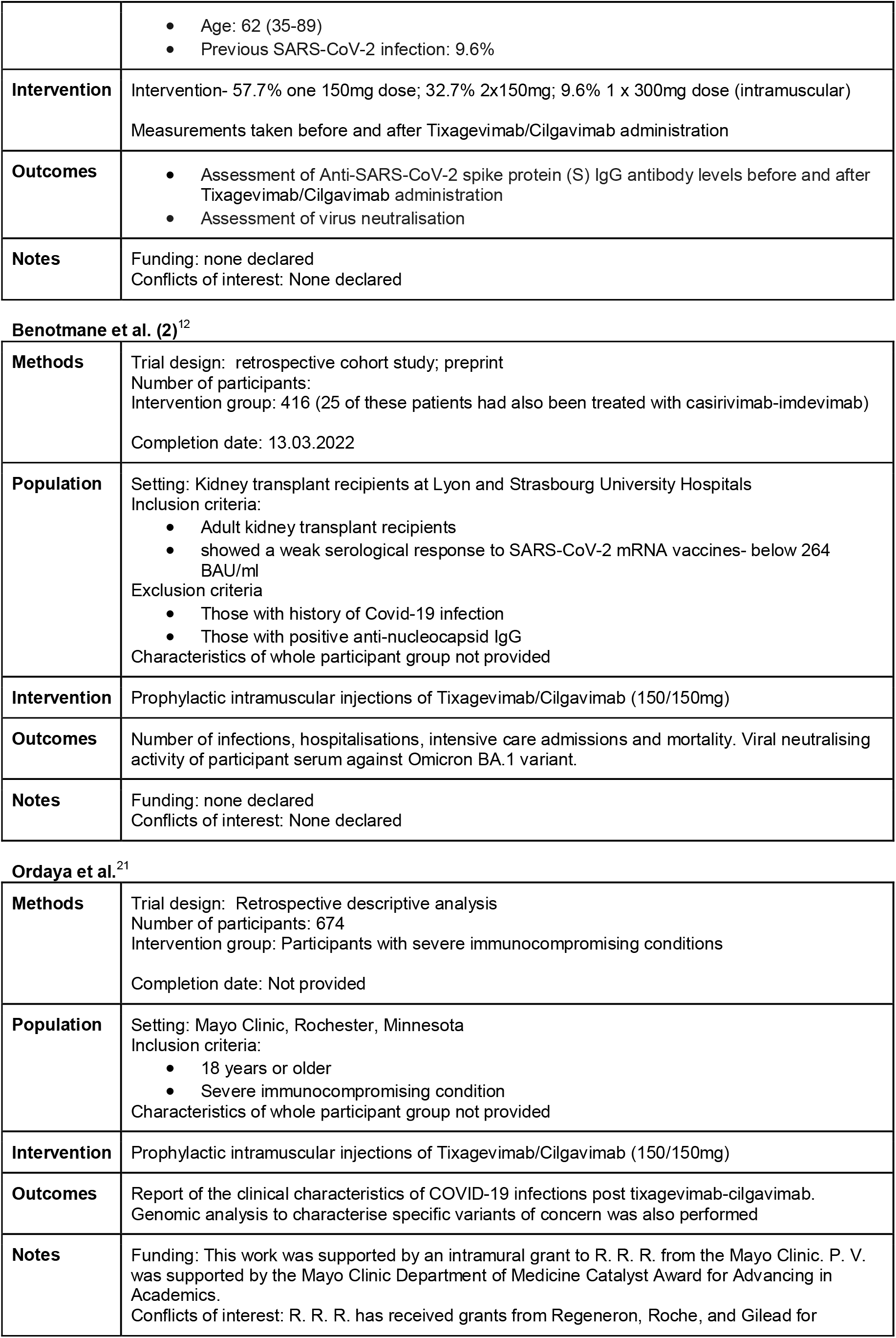
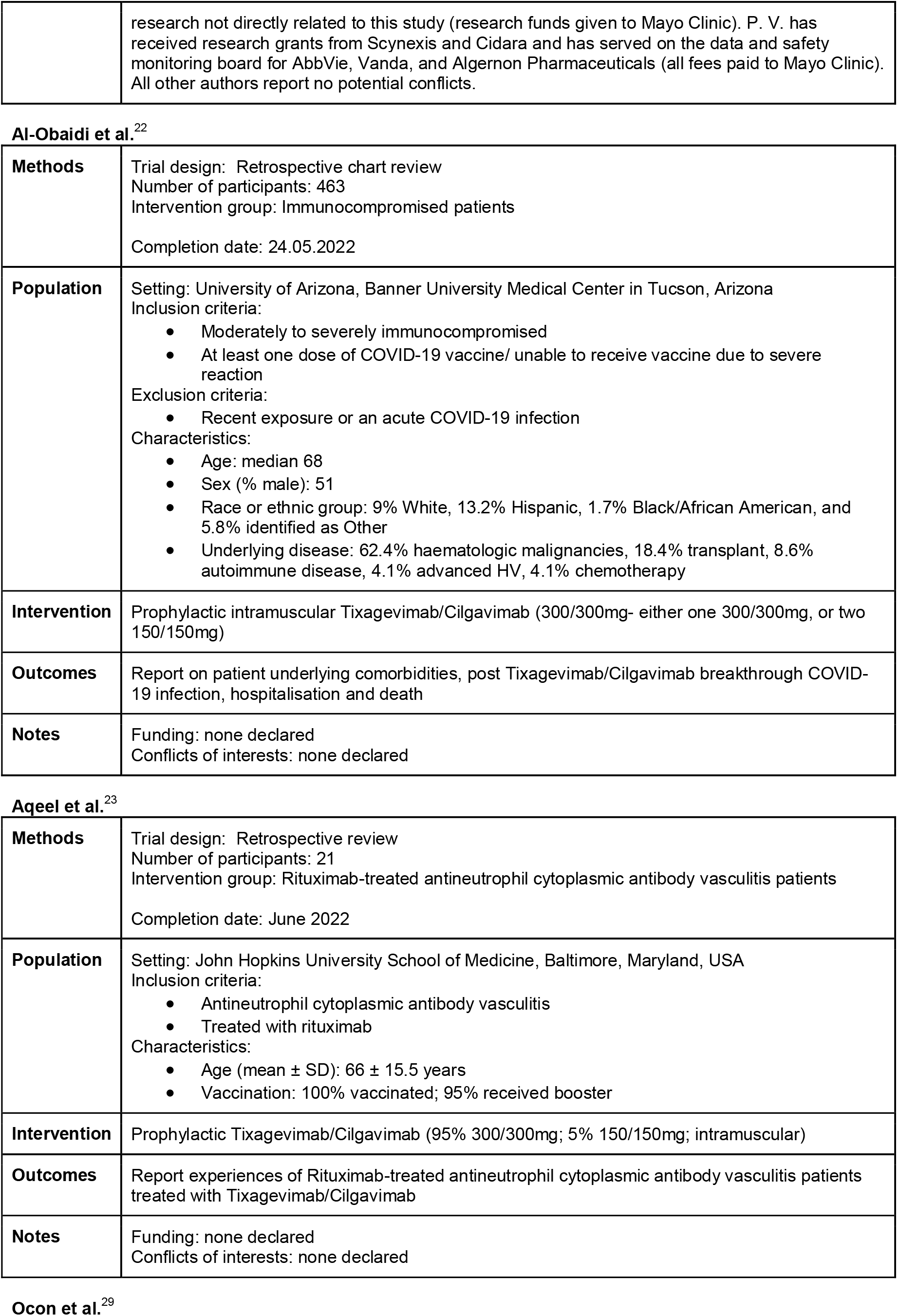
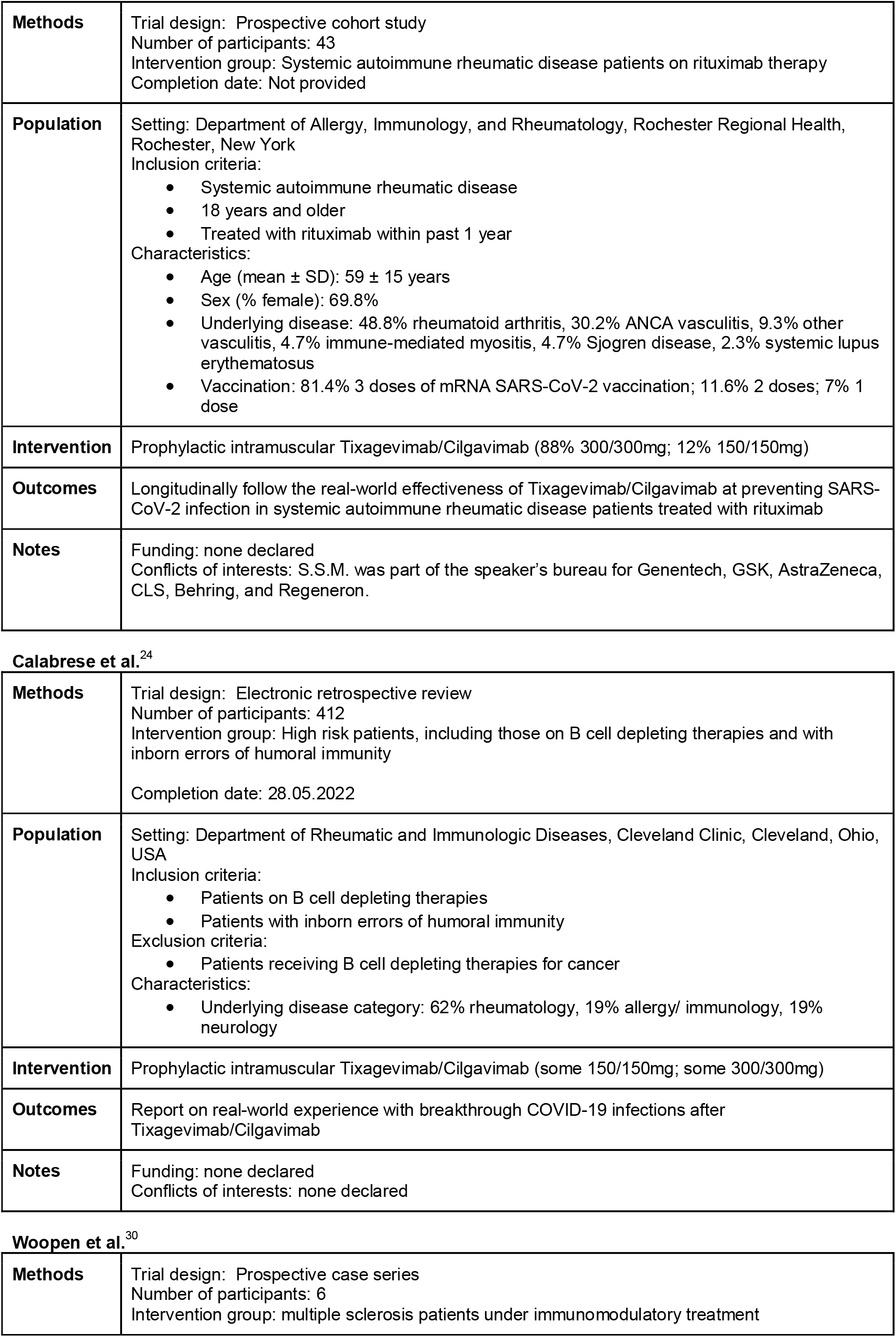
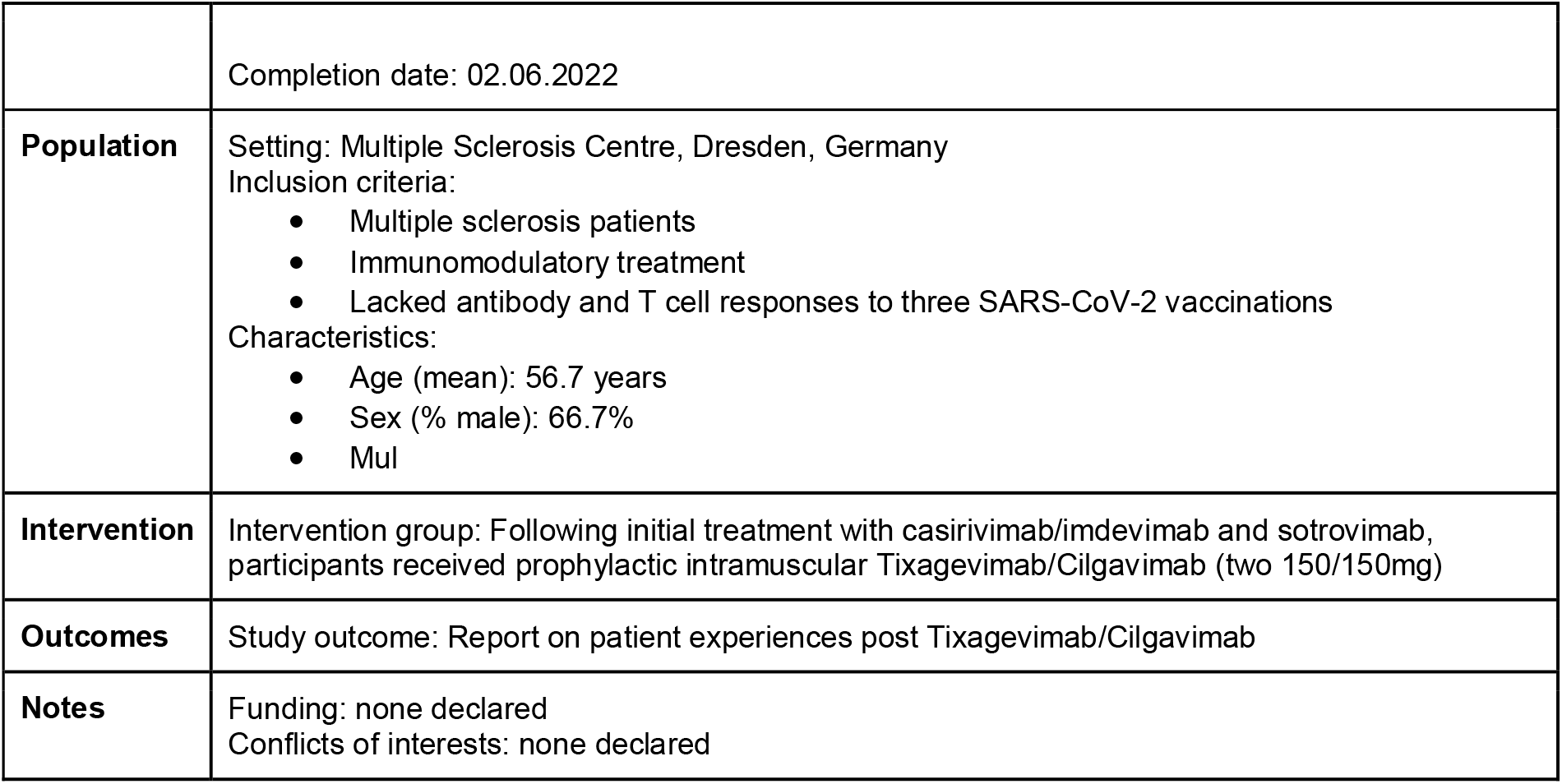

